# Mapping the landscape of lineage-specific dynamic regulation of gene expression using single-cell transcriptomics and application to genetics of complex disease

**DOI:** 10.1101/2023.10.24.23297476

**Authors:** Hanna Abe, Phillip Lin, Dan Zhou, Douglas M. Ruderfer, Eric R. Gamazon

## Abstract

Single-cell transcriptome data can provide insights into how genetic variation influences biological processes involved in human biology and disease. However, the identification of gene-level associations in distinct cell types faces several challenges, including the limited reference resource from population scale studies, data sparsity in single-cell RNA sequencing, and the complex cell-state pattern of expression within individual cell types. Here we develop genetic models of cell type specific and cell state adjusted gene expression in mid-brain neurons in the process of specializing from induced pluripotent stem cells. The resulting framework quantifies the dynamics of the genetic regulation of gene expression and estimates its cell type specificity. As an application, we show that the approach detects known and new genes associated with schizophrenia and enables insights into context-dependent disease mechanisms. We provide a genomic resource from a phenome-wide application of our models to more than 1500 phenotypes from the UK Biobank. Using longitudinal genetically determined expression, we implement a predictive causality framework, evaluating the prediction of future values of a target gene expression using prior values of a putative regulatory gene. Collectively, this work demonstrates the insights that can be gained into the molecular underpinnings of diseases by quantifying the genetic control of gene expression at single-cell resolution.

## Introduction

Recent years have seen a dramatic increase in the use of single-cell datasets to investigate biological mechanisms of complex diseases^1^. These developments offer new mechanistic insights by facilitating the study of the cellular and phenotypic consequences of genetic variants at high resolution. For example, these advances have allowed interrogation of expression quantitative trait loci (eQTL) at the cellular level^1,2,3^. One notable finding from these studies is the role of dynamic eQTLs with transient and condition-dependent effects^1,4^.We hypothesize that developing in silico models of lineage-specific dynamic regulatory effects on gene expression will extend our understanding of the molecular basis of complex diseases.

Bulk-tissue eQTL analysis such as from GTEx and eQTLGen have highlighted the tissue-specific manner by which genetic variants may regulate gene expression^4,5^. However, analyses based on these reference panels, despite their range of tissues (as in GTEx) or their sample size (as in eQTLGen), may be confounded by cellular heterogeneity. The expression differences seen at a tissue level could be attributed to either cell proportion difference or intrinsic transcriptome level difference in specific cell types^6^. This limitation can hinder the discovery of a risk gene, as its expression changes may be linked to a confounder or transient phenotype. Therefore, it is necessary to analyze gene expression in isolated cell types from a given condition to get an unbiased approach and capture context specificity^7^. This observation has prompted recent efforts to use population-scale single-cell-based eQTL analysis in different conditions^1,2,8^. Using colocalization methods, these studies have mapped disease-associated variants to cell-specific eQTLs and reported novel disease associations that were not detected at the tissue level^8,9^. However, single-variant colocalization analyses may limit the power to identify underlying mechanisms. Indeed, this approach ignores aggregate effects, which may be driving the observed condition specificity.

For highly polygenic traits, GWAS variants typically have modest phenotypic effects, hence a methodology that investigates the joint impact of multiple variants may enhance genetic association analysis^10,11^. Through aggregation, methods such as PrediXcan have demonstrated that gene expression can be imputed using local genetic variants^12,13^. In Transcriptome Wide Association Studies (TWAS), the imputed expression is used to identify disease-associated genes^12,13^. Causal inference on the gene’s effect on disease can be performed, leveraging multiple genetic variants as instrumental variables^13^.

Here we develop models of cell-type specific gene expression to further our understanding of context-dependent gene regulation. The approach leverages single-cell sequencing datasets involving diverse cell types and differentiation stages^8^. The method trains prediction models of gene expression, using genetic variants as features, to capture gene expression’s cell-type or temporal specificity. Cellular state, a transient phenotype for a given cell type, can create heterogeneity within the cell type, which can limit the detection of context-dependent eQTLs^9,14,15,16^. Therefore, we also train additional prediction models after adjusting for cell state. By applying the in silico genetic models to reference individual genotype and GWAS summary data from large-scale studies, we further identify genes and regulatory networks associated with broad range of complex disease.

## Results

### Building models of gene expression from single-cell data

We developed cell-type-specific, genetic-variation-based gene expression models in induced pluripotent stem cell (iPSC) differentiated dopaminergic neuronal cells (**Methods**)^17^. We used single-cell transcriptomes of eight different cell types differentiated across three time points. The scRNA-Seq data contains transcriptome of cells captured at day 11 (D11), day 30 (D30), and day 52 (D52). The D11 dataset contains early progenitor cell types such as floor plate progenitors and neuroblasts. At D30 and D52, the scRNA-Seq data contains more specialized midbrain neurons. For some cell types (progenitors, dopaminergic, and ependymal), we have scRNA-Seq data at multiple time points (D30 and D52 or D11 and D52), and we trained models for each time point independently. After quality control (QC), normalization, and clustering analysis, we created a pseudo-bulk transcriptome matrix by aggregating the expression profile for each gene across individual cells within a specific cell type at a specific time point (**Methods**). Cell types, with cell count ranging from 9779 to 134084, were used for downstream analysis. For individuals with matching genotype data, we trained a set of “cell type specific models” for 8 different cell types (**Methods**). To account for heterogeneity within a cell type, we also generated a second set of models, “cell state adjusted models”. A schematic diagram of the methodology is described below in Fig 1.

**Fig. 1.**
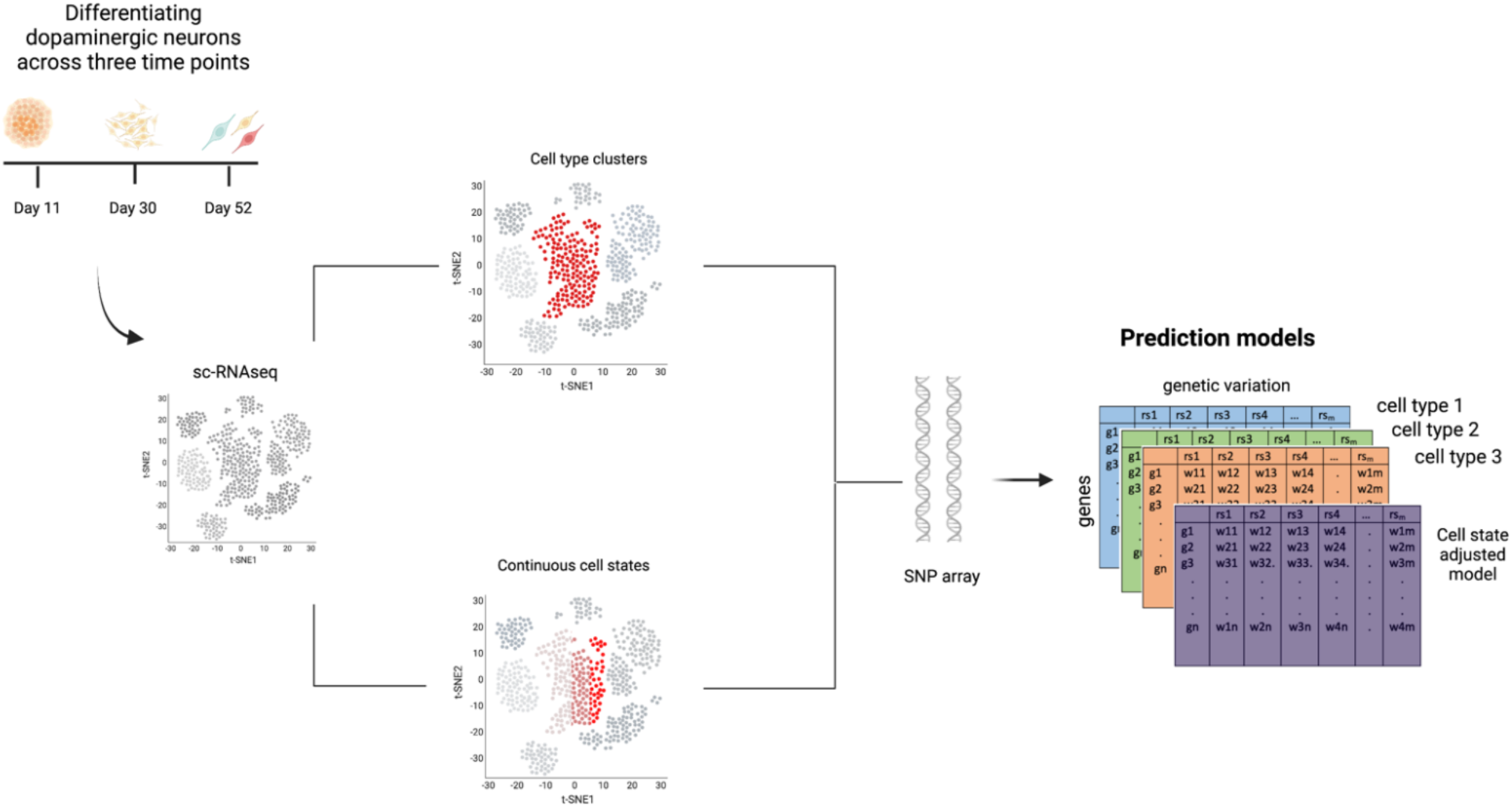
Training models of gene expression in cell types at various differentiation time points using single-cell transcriptome datasets. The workflow illustrates gene expression model building in cell types at various time points. The training dataset consists of single-cell transcriptome profiling for cells undergoing differentiation at three time points as well as matching genotype data. We created two sets of gene expression prediction models: cell type specific models and cell state adjusted models using a machine learning method. For the latter, we used principal components as covariates to adjust for continuous cell states (see **Methods**).

### Prediction performance of cell type specific and cell state adjusted models

We identified imputed genes (iGenes) across the 8 cell types and 3 time points. The number of detected iGenes varied depending on the cell type as well as the time point (Fig. 2a). The mean number of iGenes was 1486 at D11, 1398 at D30, and 1194 at D52. For the cell types with models at different time points, such as the floor plate progenitors (FPP) and proliferating floor plate progenitors (PFPP), the number of iGenes decreased (from 1662 to 1243) as the cells became more specialized. The decrease in the number of iGenes with differentiation time mirrors the decrease in the number of discovered eQTLs observed in the previous work using the same dataset^2^. We also observed a greater number of iGenes and improved performance with increased sample size (Fig 2b.). We compared the number of iGenes that are cell type specific with those genes that are shared between cell type models of the same time point. Our results show that a greater percentage of the iGenes were imputed only at specific cell types; this is evident in all time points (Supplementary Figure S1).

**Fig. 2.**
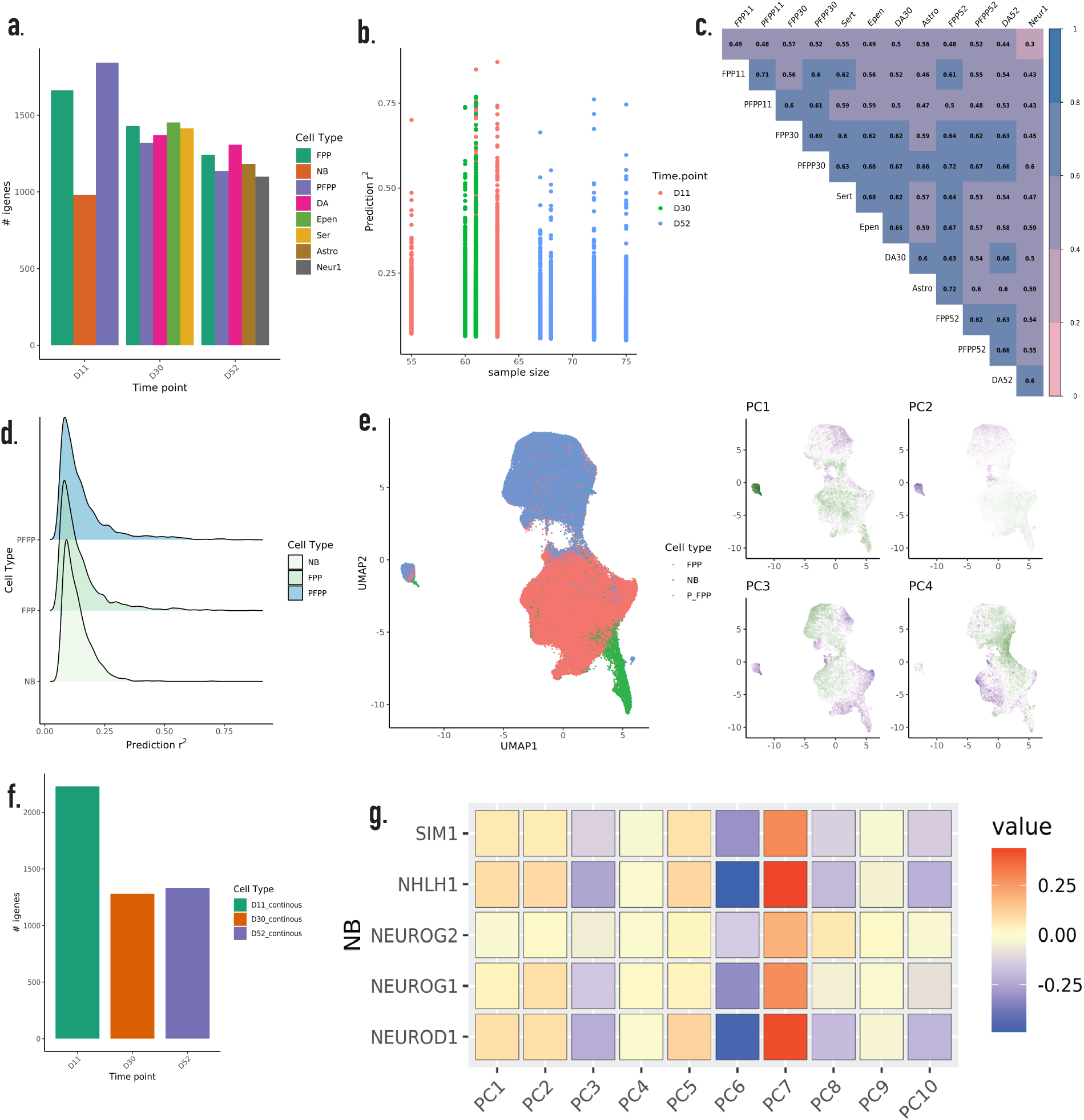
Comparison of prediction performance between cell type and cell state adjusted models. **a**, The number of imputable genes (iGenes) for the 8 cell types across three developmental time points. **b,** Sample size and prediction performance R^2^ across the three time points. **c**, Correlation of prediction R^2^ between the different cell type models. **d**, Similar empirical distribution of prediction performance observed across gene expression models in each cell type. **e,** UMAP plots of the set of dopaminergic neurons at D11 and principal components (PCs) representing continuous cell states. **g**, Correlation of calculated PCs with expression of marker genes for the Neuroblast (NB) cell types at D11. **f,** Number of iGenes for cell state adjusted models by differentiation time point. FPP = Floorplate progenitors, PFPP = Proliferating floorplate progenitors, NB= Neuroblasts, DA = dopaminergic neurons, Epen = Ependymal cells, Ser = Serotonergic cell types, Astro = Astrocyte-like cell types, Neur1 = Neurons. D11 = day11, D30 = day 30, D52 = day 52, PC = Principal Component, UMAP = Uniform Manifold Approximation and Projection

We evaluated the prediction performance (R^2^ values) of the cell type specific and the cell state adjusted models at three time points, D11, D30 and D52 (Fig. 2e). For the cell type models, we observed an average R^2^ of 0.15 at D11, 0.13 at D30, and 0.12 at D52. The median performance was 0.11 for the gene expression prediction models at D11 and D30, a median of 0.10 for D52 (Fig. 2d). We further compared the correlation of the prediction performance among the different cell type models. We observed strong correlation (rho = 0.56) between the cell type models at the same time point (Fig. 2c).

Next, we compared prediction performance between time points in the same cell type. We observed that some genes showed improved prediction performance at a specific time point (Supplementary Figure S2). We further examined the performance of early lineage cell type models (D11) in their ability to predict late gene expression (D30 or 52) in a cross-validation framework. To quantify the accuracy of the prediction, we tested the correlation of the genetically predicted expression at the earlier time point with the measured transcriptome for that cell type^18^. To account for the bias that may arise due to sample overlap, we compared the *R*^2^ calculated using the actual expression dataset and the *R*^2^ calculated using randomly selected gene expression datasets in up to 1000 permutations. Those genes with an empirical p-value of < 0.05 were deemed significantly predicted. We observed that earlier time point modes (D11 or D30) were only able to predict smaller portion of the iGenes at the later time point (D30 or D52), emphasizing that gene expression prediction is sensitive to cellular developmental stage. The significantly predicted genes also have higher baseline mean expression than the remaining genes (Supplementary Figure S9).

### Characterizing cell state adjusted models of gene expression

Aggregating cells into discrete clusters during model training imposes limitations on capturing the continuous transcriptional landscape^8,15,16,19^. Using a dimensionality reduction method, the single-cell transcriptome data can be decomposed to multiple functional cell states. Following quality control procedures, we initially computed Principal Components (PCs) to characterize cell states across all cell types at each time point. Subsequently, we accounted for the influence of the top 10 PCs, by regressing them out from the normalized expression data and utilizing the residuals for model training (Fig. 2e). By employing this approach, we were able to construct prediction models that are independent of continuous cell states. In contrast to the cell type models we developed above, these new models offer the potential to examine the influence of cell state independence on model training^9,15,16^. We observed that individual PCs correlated with lineage specific cell type marker genes (Fig. 2e), suggesting a notable relationship between cell type identity and cellular state (Fig. 2g, Supplementary Figure S4)^20^.

We analyzed the performance of the cell state adjusted models. As in the cell type specific models, the number of iGenes decreased at later time points (Fig. 2f). In addition, some genes were predicted with higher accuracy using the cell state adjusted models whereas for other genes, the performance was enhanced using the cell type models (Supplementary Figure S8). We investigated whether the improved performance from the cell state adjusted models for certain genes was due to their low expression levels in the cell type. Our analysis revealed that there was no significant association (Fisher exact p-value 0.514) between low expression (defined as the lowest 25% mean log expression) and cell state specificity, suggesting that the performance gain could not be attributed to the genes’ expression levels in the cell type. As TWAS power to detect trait associations depends on model performance (reflecting the extent to which the models capture the (cis) heritability of gene expression), we make available both sets of models for downstream applications (see Data Availability).

### Cell-type and time profile specificity of gene expression

To quantify gene expression specificity across time points, we calculated the *τ* statistic from the log normalized cell-type specific RNA sequence^21^ (**Methods**). The value of *τ* ranges between zero and one. As *τ* approaches one, gene expression becomes more highly cell type specific. As expected, we found that with differentiation, there was an increase in the *τ* value, which moved increasingly closer to one (Fig. 3, Supplementary Table 14 &15). We also compared *τ* for each gene between pairs of time points. We observed the highest shift or difference in *τ* between D11 and D30 as compared to between D30 and D52 (Fig. 3a & 3b), indicating that change in cell type specificity between time points is itself dynamic. We examined whether genes exhibiting high cell type specificity, as indicated by a high *τ* value, also displayed elevated predicted performance in the cell type models. Our findings, based on the D30 cell type prediction *R*^2^ and *τ* value, demonstrate a negative correlation between performance and cell type specificity (Fig 3e). To determine the functions of the broadly expressed genes, we performed a gene ontology enrichment analysis using the bottom ten percentile of *τ*. Eighty percent of these genes were also predicted in at least one brain tissue PrediXcan model. Moreover, these genes were enriched in basic biological processes such as cytoplasmic translation, gene expression, cellular respiration (Supplementary Figure S5). However, we also observed that some genes with high τ (for example, *WFDC3*, *RPL12*) displayed improved performance in certain cell types, strengthening the premise that context dependence is important for gene expression prediction.

**Fig. 3.**
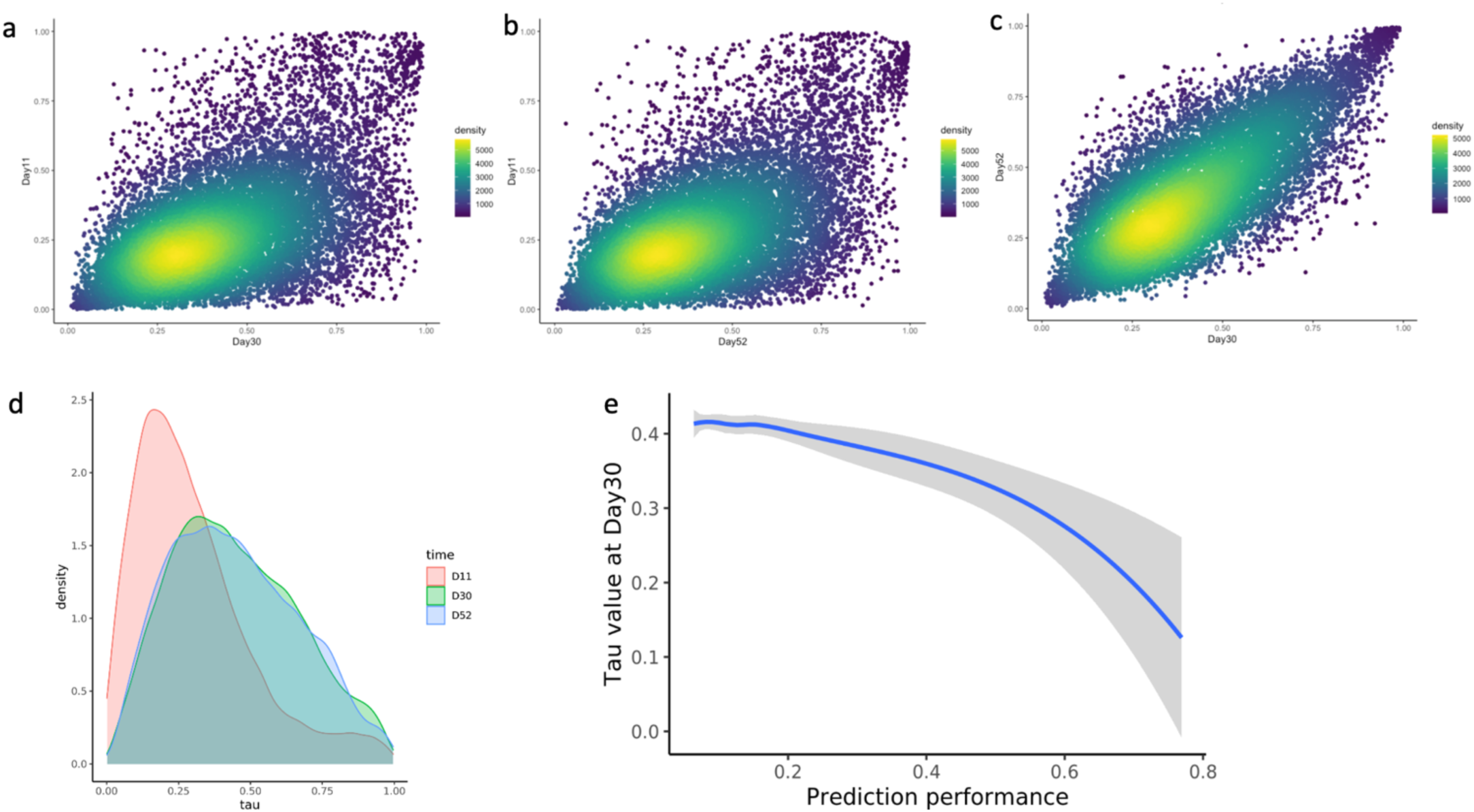
Gene expression breadth and cell type specificity across single cells at different time points. **a-c,** The tau (*τ*) index as a measurement of gene expression similarity between cell types at the three time points (D11, D30, and D52). Each dot in the scatter plot represents the *τ* value for a specific gene. A value of τ close to one shows high cell type specificity of gene expression, while a value of τ close to zero shows less specificity and wider breadth of expression across different cell types **d,** Distribution plot for *τ* values across time points. **e,** The correlation between *τ* and prediction performance R^2^ for cell type models at D30. The curve was fitted with a locally weighted smoothing (LOESS) regression.

### Application to SCZ GWAS

We applied the models of gene expression in the 8 available cell types to the latest schizophrenia (SCZ) GWAS from the Psychiatric Genomics Consortium (PGC3)^22^. We leveraged the GWAS summary statistics data to identify gene-level associations with SCZ susceptibility. Notably, we identified genes that were associated with SCZ in only one cell type as well as SCZ associated genes shared among all the cell types (Table 1). Thus, some disease associations were highly cell type-specific while some were common across all the cell types. Interestingly, we also observed differences in associations for a given cell type model at different time points (Supplementary Table 2,3), highlighting the dynamics of implicated genes.

**Table 1.** GReX associations with schizophrenia that are unique to cell type models.

| Cell type | Time point | Gene | Ensemble gene | pvalue | prediction R<sup>2</sup> |
| --- | --- | --- | --- | --- | --- |
| Floor plate progenitors (FPP) | day 11 | *ZFP57* | ENSG00000204644 | 1.41E-14 | 0.07752337 |
| Floor plate progenitors (FPP) | day 11 | *ARL17B* | ENSG00000228696 | 3.15E-11 | 0.34033268 |
| Floor plate progenitors (FPP) | day 11 | *ZNF165* | ENSG00000197279 | 2.01E-08 | 0.09104885 |
| Floor plate progenitors (FPP) | day 11 | *CHRNA3* | ENSG00000080644 | 3.15E-06 | 0.23994094 |
| Floor plate progenitors (FPP) | day 11 | *HYI* | ENSG00000178922 | 1.17E-05 | 0.34983064 |
| Neuroblasts (NB) | day 11 | *DHX16* | ENSG00000204560 | 5.07E-14 | 0.07225118 |
| Neuroblasts (NB) | day 11 | *WDR82* | ENSG00000164091 | 3.85E-06 | 0.074624159 |
| Neuroblasts (NB) | day 11 | *CS* | ENSG00000062485 | 7.29E-06 | 0.146730957 |
| Neuroblasts (NB) | day 11 | *TNXB* | ENSG00000168477 | 3.56E-05 | 0.073614859 |
| Neuroblasts (NB) | day 11 | *SMDT1* | ENSG00000183172 | 6.52E-05 | 0.1111063 |
| Proliferating FPP (PFPP) | day 11 | *BOLA1* | ENSG00000178096 | 1.80E-08 | 0.08429438 |
| Proliferating FPP (PFPP) | day 11 | *HSPA1B* | ENSG00000204388 | 1.19E-07 | 0.16794496 |
| Proliferating FPP (PFPP) | day 11 | *ZFP57* | ENSG00000204644 | 2.58E-07 | 0.10862589 |
| Proliferating FPP (PFPP) | day 11 | *SMDT1* | ENSG00000183172 | 8.38E-06 | 0.54342332 |
| Floor plate progenitors (FPP) | day 11 | *DUSP7* | ENSG00000164086 | 2.43E-05 | 0.14350724 |
| Floor plate progenitors (FPP) | day 30 | *SF3B4* | ENSG00000143368 | 1.16E-09 | 0.171576909 |
| Floor plate progenitors (FPP) | day 30 | *PITX3* | ENSG00000107859 | 2.03E-07 | 0.155739762 |
| Floor plate progenitors (FPP) | day 30 | *XPNPEP3* | ENSG00000196236 | 2.45E-06 | 0.190724139 |
| Floor plate progenitors (FPP) | day 30 | *RPL12* | ENSG00000197958 | 2.23E-05 | 0.344603902 |
| Floor plate progenitors (FPP) | day 30 | *CIAO1* | ENSG00000144021 | 2.65E-05 | 0.136599462 |
| Floor plate progenitors (FPP) | day 30 | *FAM72C* | ENSG00000203817 | 2.83E-05 | 0.111795563 |
| Proliferating FPP (PFPP) | day 30 | *GABBR1* | ENSG00000204681 | 8.99E-07 | 0.066050948 |
| Proliferating FPP (PFPP) | day 30 | *RPL13* | ENSG00000167526 | 4.63E-06 | 0.101211532 |
| dopaminergic (da) | day 30 | *MPHOSPH9* | ENSG00000051825 | 6.07E-13 | 0.25087399 |
| dopaminergic (da) | day 30 | *NELFE* | ENSG00000204356 | 6.51E-08 | 0.16189148 |
| dopaminergic (da) | day 30 | *KCNB1* | ENSG00000158445 | 6.54E-08 | 0.14616822 |
| dopaminergic (da) | day 30 | *ZNRD1* | ENSG00000066379 | 1.28E-06 | 0.06704342 |
| dopaminergic (da) | day 30 | *SHC1* | ENSG00000160691 | 2.69E-06 | 0.08372363 |
| dopaminergic (da) | day 30 | *IFT81* | ENSG00000122970 | 9.08E-06 | 0.06511382 |
| dopaminergic (da) | day 30 | *CAMKK2* | ENSG00000110931 | 1.20E-05 | 0.58603189 |
| dopaminergic (da) | day 30 | *RPL12* | ENSG00000197958 | 3.16E-05 | 0.10602019 |
| Ependymal -like | day 30 | *PHF7* | ENSG00000010318 | 7.45E-07 | 0.166881888 |
| Serotonergic | day 30 | *MPHOSPH9* | ENSG00000051825 | 7.67E-19 | 0.271484955 |
| Serotonergic | day 30 | *NT5DC2* | ENSG00000168268 | 3.91E-07 | 0.066129462 |
| Serotonergic | day 30 | *LSM2* | ENSG00000204392 | 3.25E-05 | 0.077044896 |
| Floor plate progenitors (FPP) | day 52 | *SLC39A4* | ENSG00000147804 | 4.04E-06 | 0.063752163 |
| Floor plate progenitors (FPP) | day 52 | *ZSCAN12* | ENSG00000158691 | 5.35E-06 | 0.119364051 |
| Proliferating FPP (PFPP) | day 52 | *TRIM39* | ENSG00000204599 | 1.24E-13 | 0.09216467 |
| Proliferating FPP (PFPP) | day 52 | *SFMBT1* | ENSG00000163935 | 9.87E-07 | 0.06778971 |
| Proliferating FPP (PFPP) | day 52 | *LY6G5B* | ENSG00000240053 | 4.77E-06 | 0.07762385 |
| Proliferating FPP (PFPP) | day 52 | *ECM1* | ENSG00000143369 | 5.07E-05 | 0.11326657 |
| Proliferating FPP (PFPP) | day 52 | *CRYBB2* | ENSG00000244752 | 8.08E-05 | 0.33644001 |
| dopaminergic (da) | day 52 | *HEPN1* | ENSG00000221932 | 1.03E-06 | 0.089131099 |
| dopaminergic (da) | day 52 | *ZBTB18* | ENSG00000179456 | 1.06E-06 | 0.073885592 |
| dopaminergic (da) | day 52 | *LRRC4* | ENSG00000128594 | 6.44E-06 | 0.054088373 |
| Ependymal -like | day 52 | *NPIPB13* | ENSG00000198064 | 2.57E-11 | 0.137869485 |
| Ependymal -like | day 52 | *RAP1A* | ENSG00000116473 | 3.35E-06 | 0.072117707 |
| Ependymal -like | day 52 | *ATP23* | ENSG00000166896 | 1.01E-05 | 0.099310659 |
| Ependymal -like | day 52 | *HLA-DRA* | ENSG00000204287 | 1.11E-05 | 0.110777535 |
| Ependymal -like | day 52 | *HSPA1B* | ENSG00000204388 | 1.31E-05 | 0.140574217 |
| Neurons | day 52 | *HIST1H2BN* | ENSG00000233822 | 7.10E-10 | 0.09508357 |
| Neurons | day 52 | *ADSL* | ENSG00000239900 | 9.46E-09 | 0.06582031 |
| Neurons | day 52 | *C6orf136* | ENSG00000204564 | 4.15E-05 | 0.12737249 |
| Neurons | day 52 | *DAXX* | ENSG00000204209 | 5.38E-05 | 0.10967416 |
| Astrocyte- like | day 52 | *DESI1* | ENSG00000100418 | 1.68E-08 | 0.114742637 |
| Astrocyte- like | day 52 | *NCEH1* | ENSG00000144959 | 1.69E-05 | 0.134359495 |

We asked whether each significantly associated gene in a cell type model would have been detected using the standard tissue-level PrediXcan models^12^. Out of the 76 SCZ associated genes across all cell type models, we observed that 25 were indeed also significant in the tissue models. Of the remaining genes, 37 were predicted in at least one cell type or a tissue brain model, and 14 genes were predicted only in the cell type models (Supplementary Table 2). We examined the genes that were predicted by both tissue and cell type models and assessed whether there was any improvement in prediction performance for the cell type models. Some genes did indeed show improvement in the cell type models but others had better performance at the tissue level (Supplementary Figure S3, S10). The latter would be expected, for instance, if the tissue was an imperfect proxy for a causal non-represented cell type.

We tested whether the set of genes predicted at an earlier time point displayed a greater departure from null expectation than the set of genes predicted at a later time point. We observed that the degree of departure from the null depended on cell type and time point, indicating strong context specificity of the associations of GReX with SCZ (Supplementary Figure S6).

### Gene level PheWAS of SCZ-associated genes

For the genes significantly associated with SCZ in just the cell type models (Table 1), we sought to characterize their effects on the disease phenome by performing phenome-wide association studies (PheWAS). We applied our cell-type specific and cell state adjusted models to > 1500 UK Biobank GWAS summary data to obtain gene-level associations (Fig. 4). As an example, we chose the genes that were significantly associated with SCZ and outside the MHC region (*CHRNA3*, *ARL17B*, and *HYI*) when using the model for floor plate progenitors at D11. The traits implicated by these genes in the PheWAS analysis highlight the enrichment for blood cellular traits such as erythrocyte and leukocyte count as well as red blood cell distribution width. The link between blood cell traits and SCZ has been previously noted, as well as the role of immune system in the disease etiology^23,24,25^. For another cell type model example, see Supplementary Figure S7. To enable downstream applications, we provide the full list of associations from the application of the cell type models to GWAS summaries of UKBB.

**Fig. 4.**
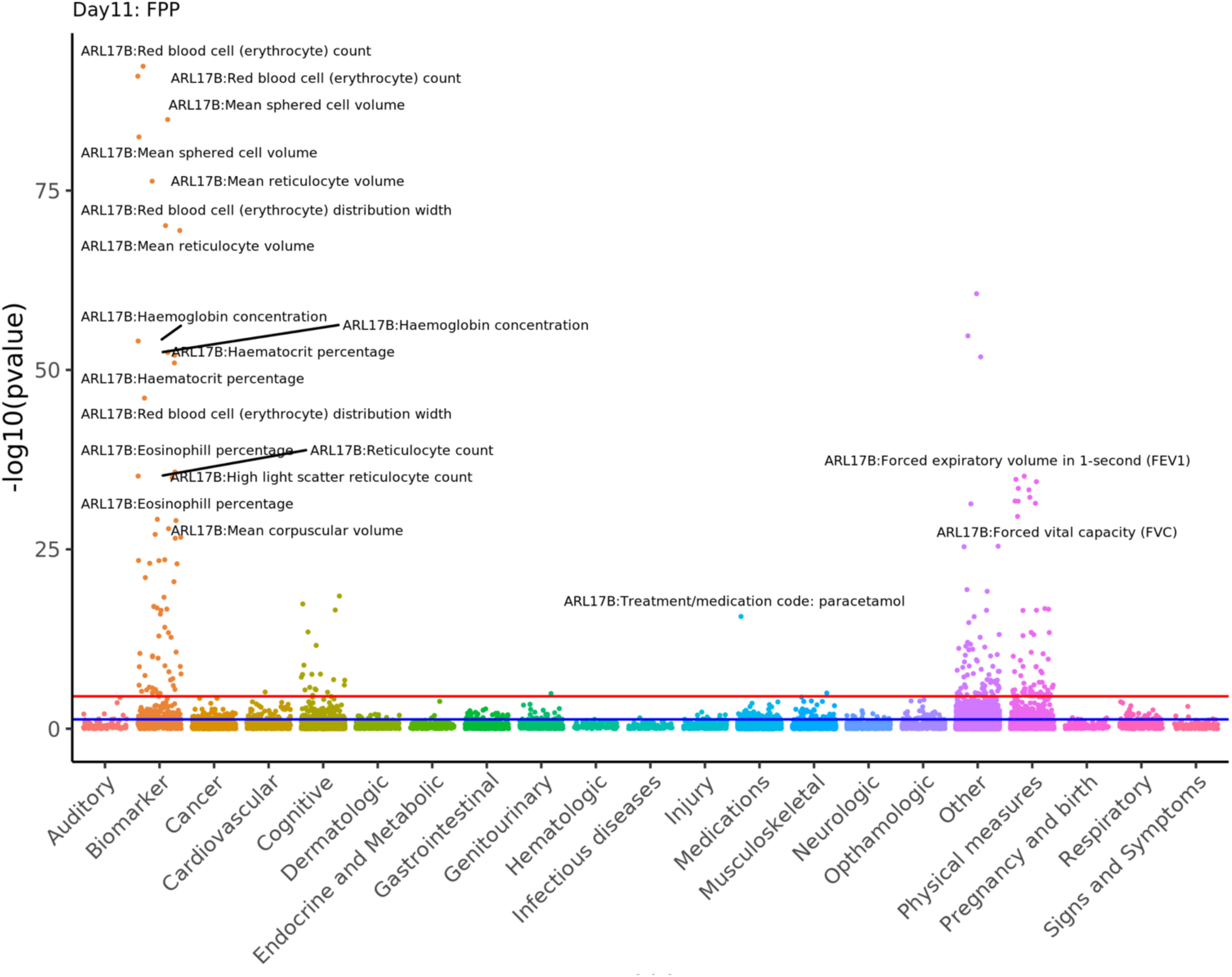
PheWAS of genes associated with SCZ using the FPP models at D11. Each data point in the Manhattan plot represents the association of a SCZ-associated gene that was significant only using the floor plate progenitors derived cell models at D11. The x-axis is the list of phenotypes grouped by category. The y-axis is the negative log10 of p-value from the association. The color coding of the data points represents disease classes, red horizontal line represents the Bonferroni threshold, and blue horizontal line represents p-value threshold of 0.05.

### Post-TWAS analysis: Identifying regulatory networks for SCZ associated genes

The identification of regulatory mechanisms that act on individual genes can provide context-specific understanding of transcriptional regulation and shed light on the role of dysregulation in disease^26–28^. Here, we sought to identify genes that are putative targets of significantly associated SCZ genes in the cell type models. Analyzing time-series data for gene networks provides a holistic view of the evolving transcriptome in complex biological systems^29^. Towards this end, we performed Granger causality inference, to test the predictive ability of the expression profile at a putative regulator at a given time point (D11 or D30) with the expression level of a hypothesized target gene at a later time point (D30 or D52) (see **Methods**). Our framework assumes the following properties of a causal gene: 1) temporal precedence (i.e., the expression of a causal gene precedes the effect [expression] on a target gene) and 2) informativeness (i.e., the expression of a causal gene harbors information about the future expression values of the target gene). Applying the cell type models to individual level reference genotype data from 1000 Genomes European-ancestry samples, we leveraged the genetically determined expression to identify regulatory genes that were Granger-causal on their targets. For significant Granger-causal pairs, we further performed motif enrichment analysis using the HOMER method (Fig. 5a) to identify a potential (transcription-factor-mediated) mechanism underlying the observed Granger-causal relationships. For illustration, we show the Granger causality output for the SCZ associated gene *HLA-DQB1* in dopaminergic cell type model at D52 (p-value = 6.25e-07) (Fig. 5b), including, at D30, *NELFE*, a subunit of the NELF complex, which negatively regulates the elongation of transcription by RNA polymerase II.

**Fig. 5.**
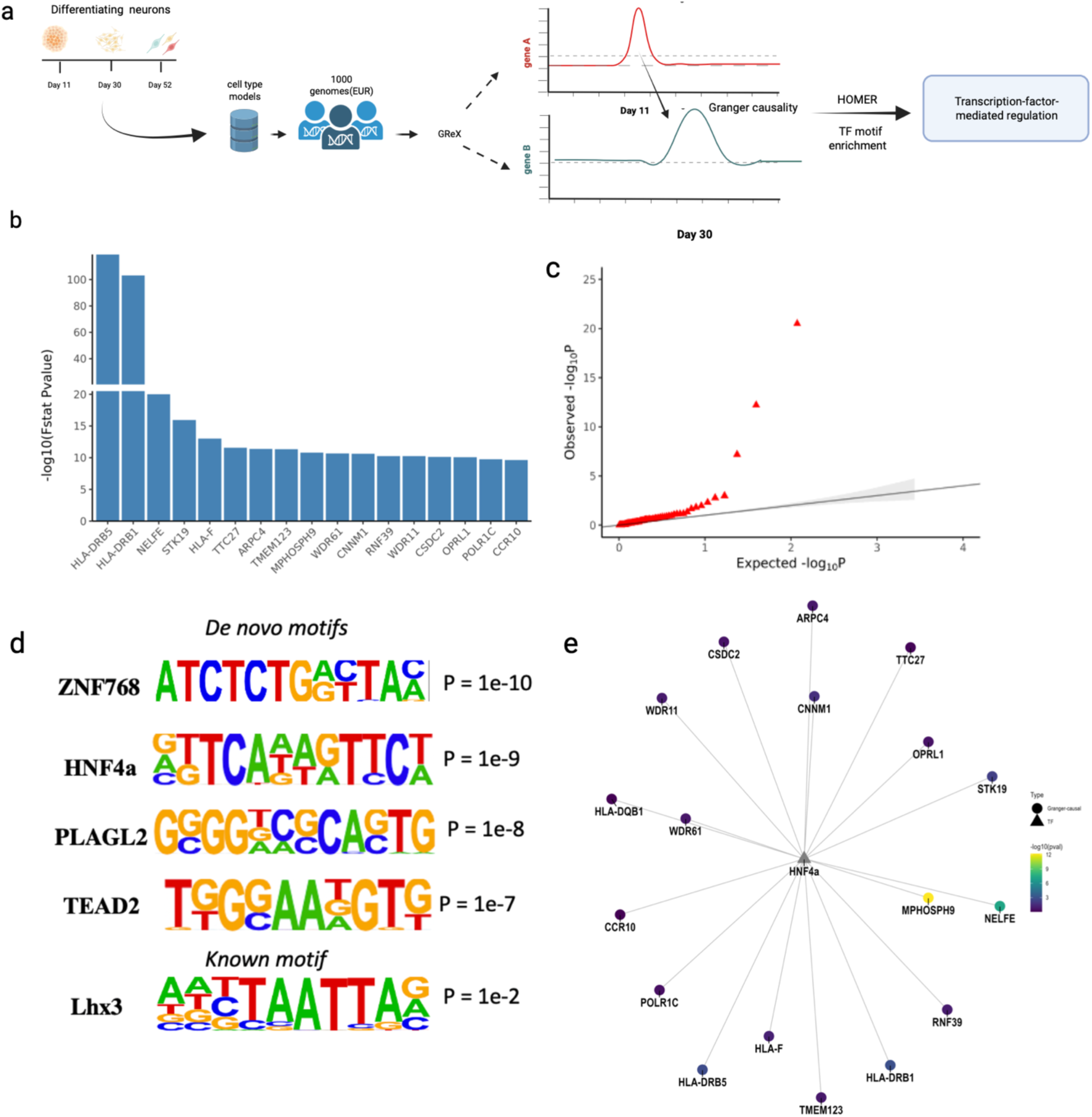
Identifying regulatory networks using Granger causality inference on longitudinal genetically determined expression data. **a,** The diagram illustrates the Granger causal inference framework for testing the effect of a putative regulatory gene on a target gene. The framework incorporates two cell type models at different time points and reference individual level genotype data. **b,** Prioritized regulatory genes obtained from the Granger causality inference for the gene **HLA-DQB1**, which is associated with schizophrenia using the cell type model for DA52. **c,** Q-Q plot showing the gene-level associations for the genes at D30 that are Granger-causal on a SCZ-associated gene at D52. **d,** Top enriched *de novo* and known transcription factor motifs for the prioritized genes found to be Granger-causal on the gene **HLA-DQB1**. **e,** Motif network showing the relationship between the transcription factor **HNF4a** and the other genes at D30, which are Granger-causal on **HLA-DQB1** at the later time point D52. Node color represents SCZ association p-value at D30.

The QQ plot of the association results for the genes from the application of the D30 models displayed a departure from the null expectation (Fig. 5c). Furthermore, to confirm whether these associations were driven by a common transcription factor (TF), we tested whether the promoters of these genes were enriched for the same motif using the HOMER method (**Methods**). Notably, our results showed an enrichment for both known and de novo motifs (Fig. 5d-e, Supplementary Figure S11). In summary, we identified a list of enriched transcription factors which can potentially regulate a network of disease relevant genes upstream. The full list of results for other cell types can be found in Supplementary Tables 16-19.

## Discussion

Our study investigated the dynamic regulation of gene expression using single-cell transcriptome data from differentiating cell types. We trained genetic models for eight distinct mid-brain neuronal cell types derived from iPSCs. Through this, we identified genes whose effects on disease risk were discernible during specific developmental stages. Our methodology enables the exploration of context-dependent genetic control of gene expression.

Despite the smaller sample size used in training the cell type models compared to the tissue models, we identified iGenes with count roughly of the same order as the number discovered using the PrediXcan tissue models. For example, the number of iGenes identified in brain hippocampus and hypothalamus (n= 81 and 82, respectively) in the earlier GTEx v6 release is 3441 and 3641, respectively; for the cell type models such as floor plate and proliferating progenitors (n=60 and n=64, respectively), the count is 1662 and 1884, respectively. This suggests that enhancing the training set sample size can lead to improved prediction in the cell type models, as also previously observed in the tissue models. Additionally, we detected many iGenes that were specific to individual cell types. By adjusting for cellular state, we were able to identify genes whose expression could not be imputed in the baseline cell type models. We also predicted more genes at an early stage, despite the lower sample size, than at a later stage; this finding is similar to that from the previous study, where more eQTLs were detected at the progenitor (earlier) cellular stage compared to later time stages. This observation could potentially be explained by our result that revealed a negative correlation between prediction performance and the cell type specificity of gene expression, as quantified by τ. As genes exhibit broader expression and possess less specialized functions in initial developmental stages, they may be more readily imputable using our model assumptions. However, we also observed that some genes with high τ (i.e., not broadly expressed) can have improved prediction performance in specific cell types or time points. This suggests that, though performance for these genes may be lower than for broadly expressed genes, leveraging the right context will improve prediction performance. In sum, these results indicate that training models using single-cell RNA sequencing data derived from cells in the process of specializing allows incorporation of dynamic eQTLs in gene prediction, identifying new iGenes that were not well predicted at the tissue level.

When applied to the SCZ GWAS data, the cell type models detected 51 significant associations that are unique to this class of models. This gain in association count can be attributed to new predicted genes (e.g., *MPHOSPH9, HEPN1, PITX3, TRIM19*). Another reason for the gain in the number of associations was improved prediction performance such as in the case of these genes (*CUL3, PHF7, LSM2, CS, WBP1L*). Generally, we observed improved prediction performance for the HLA genes in the cell type models (*HLA-C, HLA-B, HLA-DMB, HLA-DMA*). Importantly, some genes were found to be uniquely associated with SCZ at the tissue level but were neither imputed nor passed the significance threshold at the cell type level. This suggests that we may not have the relevant cell type and further highlights the need for more comprehensive single-cell transcriptome data. Overall, our approach shows that incorporation of cell type and differentiation time specific eQTLs in TWAS, can identify disease gene associations not accessible via the tissue models, hence uncovering one of the sources of missing regulation. By collectively using cell type, time point, and context specific genetic prediction models, these findings provide important insights into context-specific genetic regulation of gene expression and provide a strategy to enhance the identification of new disease associations.

We also implemented a causal inference framework, leveraging the time series single-cell transcriptomic data to identify a wider network of disease relevant genes. In contrast to previous methods such as SINGE, LEAP, and SINCERITIES, our approach does not depend on pseudo-time ordering of cells^37,38,39^. Moreover, most of these methods calculate gene correlation within each pseudo-time window, while some methods further generate the correlation matrix across time points. Here, we implemented a bivariate regression approach, to test for the Granger-causal effect of the expression of a putative regulator at an initial time point on the expression of a target gene at a later time point. Even though the number of time points is limited, we found that the type I error was well calibrated. In addition, rather than using directly measured gene expression, our approach used the genetically determined expression, which, by design, reduces the environmental confounding, as an input.^12^ By selecting genes Granger-causal on an SCZ associated gene, and subsequently conducting motif enrichment analysis using HOMER, we identified transcription factors (TFs) and co-regulated genes that may play a role in the development of the disease. Our results showed that we can capture transcription-factor-mediated regulation, opening new possibilities for connecting dynamic gene regulation with disease mechanism.

The framework presented here has certain limitations. Given the dataset’s emphasis on differentiation, the initial time point primarily consists of progenitor cells, while the subsequent time points exhibit a higher prevalence of mature neuronal cell types. It is worth noting that certain cell types have only one available time point for RNA sequencing data, leading to the training of a single time point model. Consequently, we conducted separate investigations to examine the influence of cell type and time point specificity on gene expression prediction. A more comprehensive analysis of time sensitivity would be feasible if data were accessible to track multiple cell types at multiple time points. Moreover, lack of similar datasets makes replication studies challenging. Nevertheless, we cross-validated the prediction performance as well as GWAS association results using well established tissue level models from PrediXcan. We expect that increasing sample size in similar datasets will improve prediction and facilitate improved performance. Although the models were not trained using cells directly obtained from human patients, the utilization of induced pluripotent stem cells (iPSCs) and derived neurons has proven valuable as they have been employed to identify target genes for subsequent functional validation^1,17^.

## Methods

### HipSci genotype processing

We obtained the genotype data for the human induced pluripotent cell lines (iPSCs) derived from dermal fibroblasts generated by the HipSci consortium^17^. The samples were of European ancestry. The genotype data had been imputed and phased using reference panels from the 1000 Genomes Phase 1 and rare variants from UK10K. Individual-level genotype data were retrieved through the European Nucleotide Archive (ENA)^17^. Single-sample VCF files were merged using BCFtools and subsequent QC was performed using plink v.1.9. SNP-level. Using the *--geno* and *--mind* functions in plink respectively, we removed SNPs and individuals that are missing in a large proportion of the data. Variants and individuals with missing genotype data were removed. Moreover, variants were filtered based on the minor allele frequency (MAF) threshold of 0.05 and Hardy-Weinberg equilibrium (HWE) value of 1e-6^30^. Only autosomal variants were chosen for the subsequent steps. Around 6,505,755 variants passed quality filters and were used as input for training models.

### Single-cell data preprocessing and model training

For the single-cell datasets, processed 10x genomics data for cell clusters of each of the time points were downloaded^2^. The processed data were normalized to the total number of counts per cell and cell clusters were annotated based on marker genes manually curated from literature. We used the scanpy python package (version 1.4) for downstream analysis steps and QC of cell by gene matrices^31^. We followed QC steps as outlined in the Theis protocol^32^. We filtered out dying cells based on their mitochondrial fraction level (> 20%) and gene counts lower than 2000 and removed cells with lower than 500 genes expressed. For the cells that passed QC, we used counts per million (CPM) to normalize and log transformed the data. To prepare the pseudo-bulk gene by sample matrices, we aggregated the gene expression measures for each cell type at a given time point, across individuals, following the recently published report by Cuomo *et al*. (2020) that shows that mean aggregation results in the discovery of a maximal number of eQTLs relative to other methods such as median and sum^33^. We applied quantile normalization and used PEER to adjust for hidden covariates^34^. Following the GTEx eQTL recommendation to use 15 PEER factors for sample size less than 100, we performed PEER to adjust for hidden covariates^34^. We used the residual expression data to train the cell type specific models. We applied elastic net machine learning implemented in the R package glmnet^12^, on matching genotype and normalized gene expression data for each cell type to train the gene expression prediction models. The SNPs located within 1Mb distance of a gene’s transcription start site were used as potential features in the model training.

### Modeling cell state adjusted models

To train models that account for transitional cellular states, we extracted the scaled gene expression for cells at each time point. Following *Nathan et al. (2022)* and *Gupta et al,* (2023), we first calculated the top 10 PCs using the scaled expression^8,16^. We then regressed out the effect of the 10 PCs, treating them as covariates against the scaled expression in linear regression analysis^8,15,16^. We used the residual expression output to generate a pseudo bulk data matrix by aggregating cells for individuals. We further quantile normalized and adjusted for unknown covariates using PEER^34^. We used the resulting data as input for training models that are agnostic of cell state for each time point.

### Measuring gene expression specificity in cell types

To quantify gene expression similarity across cell types in specific time points, we used the τ index ^21^:

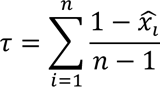

where,

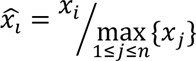

And *x_i_* is the expression level of the gene in tissue or cell type *i* and *n* is the number of cell types. For this analysis, the aggregated sc RNAseq data was normalized by CPM and log transformed. Genes not expressed in any of the cells at a time point were excluded. The τ index was then calculated for the cell types for each time point. When τ is zero, the gene is expressed similarly across the cell types during the time point. When τ is one, the gene is expressed in only a particular cell type^21^. We calculated the Pearson correlation between prediction performance and the τ value.

### Cell type and bulk tissue TWAS of schizophrenia GWAS

We downloaded the GWAS summary statistics dataset for SCZ from the Psychiatrics Genomics Consortium (PGC) and applied the models from the current study^35^. Using the summary-based S-PrediXcan approach, we also applied the 13-brain tissue (GTeX) PrediXcan models to the GWAS summary data and compared the tissue-level results and the cell-type level results^36^. We chose significantly associated genes based on Bonferroni correction of (0.05/#of gene in model in each cell type). In addition, the QQ plot of association p-values was generated for each cell type with multiple time points. Moreover, the effect size of select genes was compared across cell types.

### Gene level PheWAS of schizophrenia-associated genes

For the genes significantly associated with SCZ in the cell type models only, we sought to characterize their broad effects on the disease phenome by performing phenome-wide association (PheWAS). We applied the cell type specific models to > 1000 UK Biobank GWAS summary data using S-PrediXcan to obtain gene-level associations. We present the PheWAS results in Zenodo. To facilitate investigation of the phenome wide associations for the imputed genes, we further developed a complementary software package called “sctwas”. Leveraging the cell type model application to the UKBB GWAS data, the package allows users to investigate the list of phenome wide associations for a set of genes.

### Causal inference framework in single-cell time series

Our framework trains models of gene expression in cell types using single-cell transcriptomic measurements; these models are then applied to GWAS data to identify trait-associated genes. The framework has all the benefits of TWAS and may be extended via Mendelian Randomization, as previously implemented in JTI, to utilize genetic instruments to evaluate the causal effect of a gene (seen as exposure) on an outcome^13^.

Here we develop a complementary “causal” inference framework, leveraging the time series single-cell transcriptomic data. A causal gene on a target gene is characterized by a) temporal precedence (whereby the causal gene is prior to the effect on the target gene) and b) informativeness (whereby the causal gene holds information on the future values of the target gene). Let *g* be a causal gene for the target gene *y*. Let *g_t_* and *y_t_* be their expression level at the “current” time point *t*. A univariate model of *y* formulates the expression level *y_t_* in terms of the expression level *y_s_* from *n* previous time points, 1 ≤ *s* ≤ *n*, plus a residual term *δ_t_* at time point

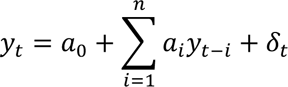

Here, for each time point *i* (where 1 ≤ *i* ≤ *n*), *a_i_* is the coefficient representing the marginal effect of *y* at that given time point on *y* at the current time point. Next, we model the effect of the causal gene *g* as follows:

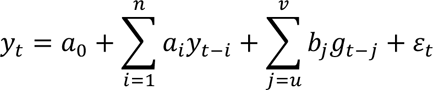

We note that this model explicitly implements the two conditions of temporal precedence and informativeness described above. When these conditions are met, we say *g* is Granger-causal on *y*. In the model, the expression level of the causal gene *g* at a previous time point *t* − *j*, where *u* ≤ *j* ≤ *v*, determines the expression level *y_t_*, with marginal effect *b_j_*. If the marginal effect *b_j_* is nonzero, for some time point *j*, then *g* is Granger-causal on *y*. We test the null hypothesis of Granger causality that the lagged values of *g* do not contribute to the variation in *y*, i.e., *b_j_* ≠ 0 for each *j*.

To perform Granger causality inference on genetically determined expression, we first applied the cell type models to individual level genotype data from the European population of 1000 genomes (n=496). We restricted this analysis to the cell types that have multiple time points such as FPP, EPEN, DA, and PFPP. Using the bivariate regression approach mentioned above, we calculated the F-stat value for each gene pair across time points to identify significantly correlated genes.

### Motif enrichment using HOMER

The Granger causality framework can be used to generate a graph *G* = (*V*, *E*), where *V* is the set of vertices and *E* is the set of edges, from the single-cell time series transcriptomic data. The graph structure encodes the functional (Granger causality derived) relationships between pairs of genes, which are represented by the adjacency matrix *A* = [*A_i_*_,*j*_]:

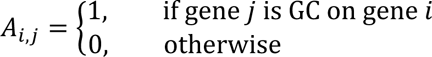

We examined whether Granger causality of a potential regulatory gene on a SCZ-associated gene could be explained by transcription-factor-mediated regulation. In effect, for SCZ-associated genes (represented as a subset of the rows of the adjacency matrix *A*), we considered the set of potential regulators (represented as a subset of the columns of *A*) with *A_i_*_,*j*_ = 1. We obtained preconfigured human promoter sets from the HOMER website (http://homer.ucsd.edu/homer/index.html). Using the command line function *findMotifs.pl*, we searched for motifs enriched in the putative target genes. This tool performs promoter based-motif analysis for both *de novo* and known TFs.

## Data availability

The cell type and cell state adjusted prediction models are available on Zenodo at (to be provided up on publication). Single-cell transcriptome data across dopaminergic neuron differentiation can be found on Zenodo (https://zenodo.org/record/4333872). The individual level genotype data are downloadable from the Human Induced Pluripotent Stem Cells Initiative (HipSci) website (https://www.hipsci.org/data). The summary statistics for the PGC data are available on the consortium’s online data repository (https://pgc.unc.edu/for-researchers/download-results/). Summary statistics for the UKBB GWAS are available at the Neale Lab online data repository (http://www.nealelab.is/uk-biobank). The tissue-specific PrediXcan gene expression models leveraged here are available for download from the JTI repository (https://doi.org/10.5281/zenodo.3842289). Phased individual level genotype data from the 1000 Genomes project can be downloaded from (https://www.internationalgenome.org).

## Code availability

All code for the QC of single-cell sequencing data set, model training, and reproduction of figures is hosted at our GitHub repository (https://github.com/gamazonlab/SingleCellPrediXcan). The code for performing transcription factor motif enrichment is available at the HOMER website (http://homer.ucsd.edu/homer/motif/). We also provide a complementary R package to allow phenome wide investigation of gene associaitons from the application of cell type models to UkBB GWAS summary statistics. The latest version of the package and details can be found in the github link mentioned above. Work-flow diagrams are created with **BioRender.com**.

## Acknowledgment

This study was supported by the following National Institutes of Health (NIH) grants to E.R.G.: NHGRI R35HG010718, NHGRI R01HG011138, NIA AG068026, NIGMS R01GM140287, and NIMH R01MH126459.

## Contributions

HA conducted the single-cell and genotype QC as well as model training. DZ assisted in the model training. PL conducted the association analysis with GWAS summary data from UKBB. DR worked on editing manuscript, interpretation of results, and provided feedback on methodology. EG supervised in designing research plan, data analysis, result interpretation, and writing of the manuscript. All authors read and approved the final manuscript.

